# Plasma proteome signatures are predictive of mortality in sickle cell disease

**DOI:** 10.1101/2025.11.04.25339477

**Authors:** Arnaud Chignon, Yosr Zaouali, Frédéric Galactéros, Pablo Bartolucci, Guillaume Lettre

## Abstract

Sickle cell disease (SCD) is one of the most common monogenic diseases in the world. This blood disorder damages all organs and is associated with severe systemic complications and increased mortality risk. Predicting SCD severity is currently difficult due to a lack of biomarkers. Here, we measured 5,411 plasma proteins in 376 SCD patients and 103 non-SCD participants to find new predictors of SCD mortality. We used protein signatures of mortality that were developed in non-SCD populations to calculate predicted mortality risk scores in our SCD dataset. The mortality scores were higher in SCD patients than non-SCD participants (P-value=3.7×10^-10^) and were associated with increased mortality in SCD patients (risk factors-adjusted hazard ratio [HR] and 95% confidence interval=2.2 [1.3-3.6], P-value=0.0032). The mortality scores correlated with several clinical variables (e.g. white blood cell count, hemoglobin concentration) and complications (e.g. leg ulcers, stroke) that are clinically relevant yet insufficient individually to predict SCD mortality. In addition to the protein signatures, we found 499 plasma proteins that associate with mortality in SCD patients (false discovery rate ≤5%), including many proteins involved in inflammatory responses such as the IL18 signaling cascade (IL18R1, IL18BP, IL18). Finally, we estimated biological age in SCD patients and non-SCD participants using the plasma proteome data. We confirmed that SCD patients age prematurely (+6.0±5.4 years older than their chronological age) and found that brain biological age positively associates with past occurrences of stroke. Altogether, our results support the use of the plasma proteome to monitor and predict clinical severity in SCD.

## INTRODUCTION

Sickle cell disease (SCD) is a recessive hematological disorder caused by mutations in the β-globin gene. It affects ∼8M individuals worldwide, mostly in Sub-Sahara Africa and the Indian sub-continent^1^. SCD is characterized by acute and chronic multi-organ complications of variable penetrance and expressivity (e.g. painful crises, stroke, neurocognitive impairment, renal failure, pulmonary hypertension)^2^, a poorer quality of life and a life-expectancy that is 20-30 years shorter than for non-SCD individuals^3^. In the United States, a recent study based on mortality data from the National Center for Health Statistics (2008-2023) showed that the mortality rate for SCD is increasing^4^. Many biomarkers have been proposed to predict SCD-related complications and mortality, although their utility remains limited due to poor statistical performance, lack of validation or challenges associated with their implementation in the clinic^5,6^.

Omics research provides new opportunities to understand clinical severity in SCD, expand the list of biomarkers that can predict SCD-related complications and prioritize new drug targets. As an example, human genetic studies of the regulation of fetal hemoglobin (HbF) levels identified the transcription factor BCL11A and paved the way to an approved gene therapy for SCD^7,8^. The development of affinity-based methods that can accurately measure 1000s of proteins simultaneously has recently enabled large-scale proteomic studies^9,10^. When combined with well-phenotyped and prospectively followed large human cohorts, these untargeted proteomic approaches can pinpoint unanticipated individual proteins or groups of proteins (i.e. protein signatures) that are predictive of human diseases. For instance, using large plasma proteome datasets available in the UK Biobank (UKBB) and other cohorts, investigators have developed and optimized protein signatures that predict chronic diseases, mortality and biological age^11–17^. Because many of the proteins detected in the plasma likely originate from a specific tissue or organ, it has even been possible to generate organ-specific predictive models of mortality and biological age from profiles of the plasma proteome^11,14^.

Previous proteomic studies in SCD have used mass spectrometry in small patient cohorts (N=15-51), but there have been few follow-up studies^18–20^. Here, we used the Olink technology to measure 5,411 proteins in the plasma of 479 participants from the GEN-MOD cohort, including 376 patients with SCD (HbSS), 51 individuals with sickle cell trait (HbAS, non-SCD) and 52 individuals without the sickle cell mutation (HbAA, non-SCD). We validated protein signatures of mortality and biological age in SCD patients, and explored how the proteomic data associate with baseline clinical variables and complications. Our results support an expanded role for the study of the plasma proteome in SCD research.

## RESULTS

### Profiling the plasma proteome of SCD patients

To identify new biomarkers of SCD-related mortality and complications, we measured 5,411 plasma proteins in participants from GEN-MOD, a prospective cohort of African-ancestry SCD patients and their relatives recruited at Hôpital Henri-Mondor (Paris, France)^21^. We included in our proteomic study 376 HbSS, 51 HbAS and 52 HbAA GEN-MOD participants (**Supplementary Table 1**). As expected, the β-globin genotype influenced the structure of the proteomic data, with the SCD (HbSS) patients separating from the non-SCD (HbAS+HbAA) participants along the first principal component (**Supplementary Fig. 1**). We found 598 proteins that were differentially expressed between SCD and non-SCD participants (|log_2_(fold-change)| ≥1 and false discovery rate [FDR] ≤5%), including many proteins that are highly relevant to SCD biology, hemolysis and stress erythropoiesis (e.g. HMOX1, HBG2, TFRC, EPO, AHSP, CAT)(**Supplementary Fig. 2** and **Supplementary Table 2**). We replicated findings from a recent study that targeted 92 proteins in 57 SCD patients and 13 healthy controls (98% validation rate)(**Supplementary Table 2**)^22^, and confirmed three proteins (HMOX1, AHSP, UMOD) that are also differently expressed between HbAS and HbAA participants (**Supplementary Table 3**)^23^. Overall, these results validate the quality of our proteomic dataset.

### Protein signatures predict SCD mortality

During a mean ∼12-year follow-up period, the survival rate among the GEN-MOD HbSS probands was 87% (40 events, 310 participants)(**Supplementary Table 1**). We tested whether existing protein signatures of mortality and aging, developed in non-SCD cohorts, could be good prognostic biomarkers in SCD. We calculated Conventional (i.e. organismal, that is all plasma proteins available) and organ-specific mortality and biological age predictions in GEN-MOD using the models developed by Goeminne et al.^11^ in the UKBB and our Olink plasma proteomic data. For downstream analyses, we only kept models that show good performance to predict chronological age, as previously reported (Pearson’s *r* >0.3) (**Methods** and **Supplementary Table 4**)^11^.

The predicted mortality risk by the Conventional model was positively associated with chronological age in both SCD patients and non-SCD participants from GEN-MOD (**Fig. 1A**). It was also significantly higher in SCD patients than in non-SCD participants (**Fig. 1B**). In SCD patients, we found that the predicted mortality risk calculated with the Conventional model was associated with mortality in GEN-MOD (hazards ratio [HR]=1.7, P-value=0.013), and this association was stronger when we corrected for known SCD mortality risk factors such as baseline white blood cell (WBC) counts, HbF levels, estimated glomerular filtration rate (eGFR) and acute chest syndrome (HR=2.2, P-value=0.0032) (**Fig. 1C**)^24^. SCD patients with a predicted mortality score in the first quartile (Q1) of the distribution were more likely to survive than patients in the fourth quartile (Q4)(log-rank P-value=0.016, **Fig. 1D**). After 5, 10 and 15 years of follow-up, the survival probability was 100%, 96.2% and 91.2% in the Q1 group, and 93.6%, 84.6% and 70.3% in the Q4 group, respectively (**Fig. 1D**).

**Figure 1.**
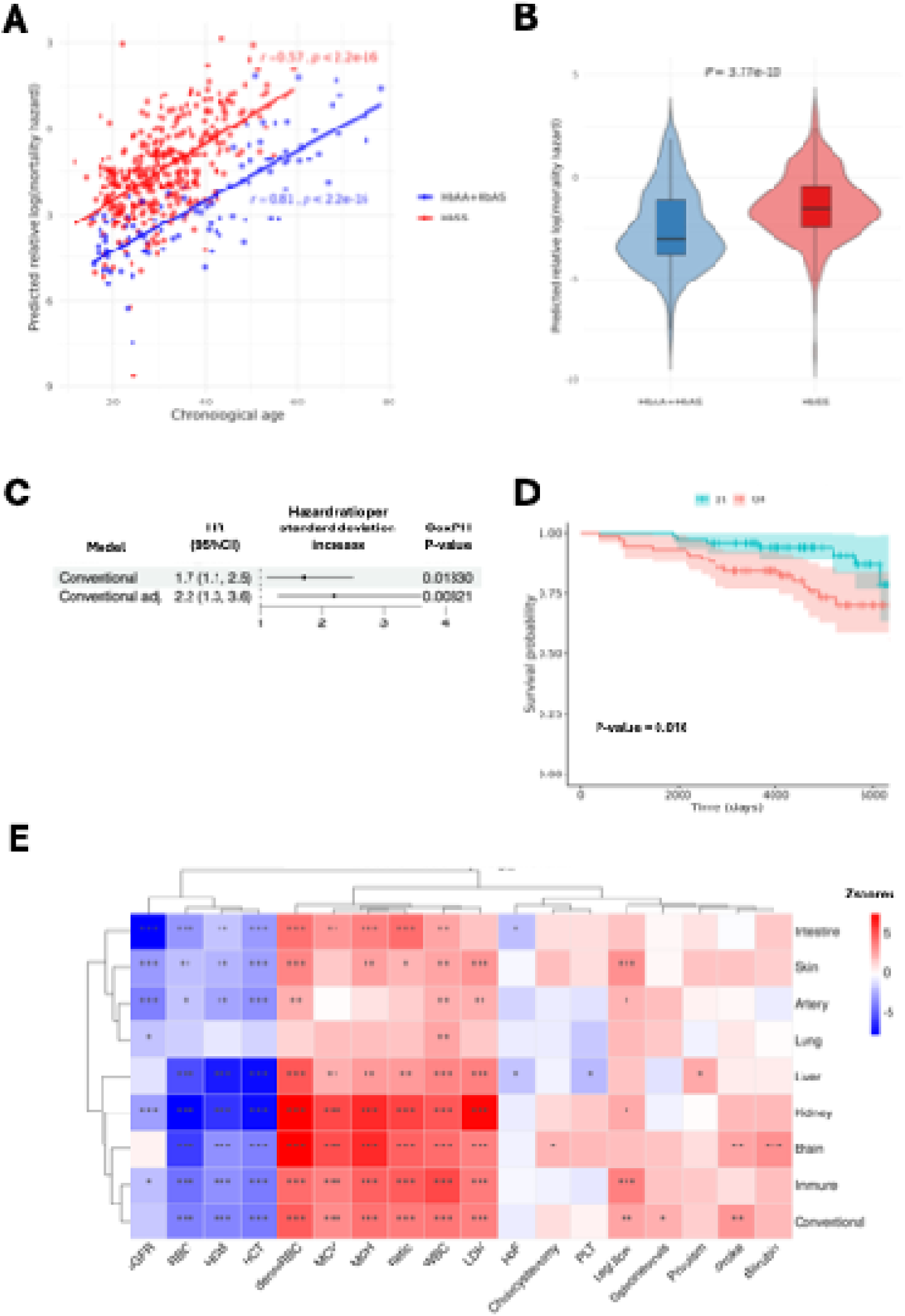
Plasma protein signatures predict SCD mortality. (**A**) Mortality predictions based on the Conventional plasma protein model increase with chronological age in SCD (HbSS) and non-SCD (HbAA+HbAS) GEN-MOD participants. (**B**) The mortality prediction scores from the Conventional model are higher in SCD patients than non-SCD participants. (**C**) The Conventional model predicts mortality in SCD patients. The two models are adjusted for biological sex and age at baseline; the adjusted Conventional model is further adjusted for baseline WBC counts, HbF levels, eGFR and acute chest syndrome. The forest plot shows the hazard ratio per standard deviation increase in the normalized prediction scores. (**D**) Kaplan-Meir plot comparing the survival probability in SCD patients with mortality prediction scores from the Conventional model in the bottom (Q1) or top (Q4) quartile of the distribution. (**E**) Heatmap that summarizes the association between mortality predictions from the Conventional or organ-specific models and baseline clinical variables. (*, nominal P-value <0.05; **false discovery rate <0.05; ***Bonferroni-corrected P-value <0.05; correction applied separately for each model). HR, hazard ration; CI, confidence interval; Cox PH, Cox Proportional-Hazards; WBC, white blood cell; PLT, platelet; Retic, reticulocyte; LDH, lactate dehydrogenase; denseRBC, dense red blood cell; MCH, mean corpuscular hemoglobin; MCV, mean corpuscular volume; RBC, red blood cell; HGB, hemoglobin; HCT, hematocrit; HbF, fetal hemoglobin; eGFR, estimated glomerular filtration rate.

The mortality prediction scores by the Conventional model were associated with many clinical variables measured at baseline: positively with dense red blood cell (RBC), mean corpuscular hemoglobin (MCH), mean corpuscular volume (MCV), reticulocyte counts, WBC counts and lactate dehydrogenase levels, and negatively with red blood cell (RBC) counts, hemoglobin levels and hematocrit (**Fig. 1E** and **Supplementary Table 5**). SCD patients with leg ulcers, osteonecrosis or stroke also had higher mortality prediction scores based on the Conventional model (**Fig. 1E** and **Supplementary Table 5**). However, none of these baseline variables were individually associated with mortality in GEN-MOD, indicating that the plasma protein signature from the Conventional model captures more information than routinely measured clinical biomarkers (**Supplementary Table 6** and **Supplementary Fig. 3**).

We explored if the organ-specific mortality predictions added additional information to the Conventional mortality model. Except for the Artery model, the mortality scores for all other organ models were higher in SCD patients than non-SCD participants (**Supplementary Fig. 4A**). The Lung, Skin and Artery models were associated with increased mortality in GEN-MOD (**Supplementary Fig. 4B**). When testing against baseline clinical variables and SCD-related complications, we found that the organ-specific mortality predictions were redundant with the mortality scores by the Conventional model, with some noteworthy exceptions: (1) eGFR was correlated with mortality scores from many organ-specific models, including the Kidney and Intestine models, (2) platelet (PLT) counts and bilirubin levels were associated with predictions from the Liver and Brain models, respectively, and (3) the presence of leg ulcers was more strongly associated with predictions from the Skin and Immune models than the Conventional model (**Fig. 1E** and **Supplementary Table 5**).

### 499 proteins associate with mortality in SCD patients

Next, we tested the association between single proteins and mortality. Using Cox Proportional-Hazards regression, we identified 499 proteins associated with mortality in GEN-MOD at an FDR ≤5% (**Fig. 2A** and **Supplementary Table 7**). Only two proteins, CBLN4 and SIRT1, were associated with increased survival. Of the 497 proteins associated with increased mortality, many are involved in inflammatory responses, including three members of the IL18 signaling cascade (IL18R1, IL18BP, IL18)(**Supplementary Table 7**)^25,26^. Using a recently released proteomic atlas that assigns plasma proteins to their tissue (or cell-type) of origin^27^, we determined that proteins associated with mortality in GEN-MOD are enriched for proteins common to most tissues (odds ratio [OR]=1.8, Fisher’s exact test P-value=2.0×10^-6^) and proteins from the spleen (OR=1.8, P-value=0.0013), and are depleted for proteins from the brain (OR=0.4, P-value=7.3×10^-6^)(**Supplementary Table 8**).

**Figure 2.**
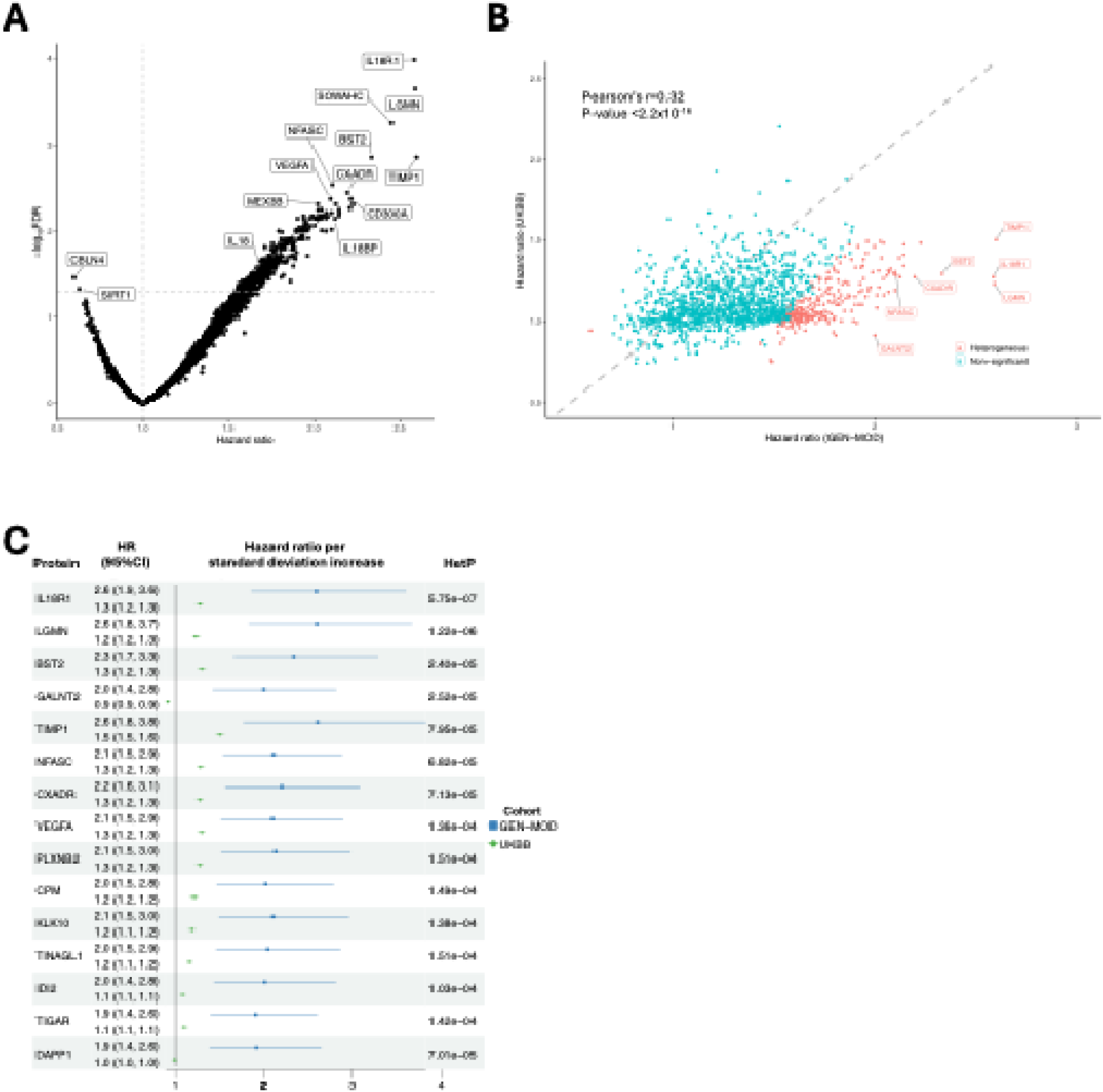
Plasma proteins associate with mortality in GEN-MOD SCD (HbSS) patients. (**A**) Volcano plot with 5,411 plasma proteins tested against mortality in SCD patients from GEN-MOD. We found 499 proteins that are significant after accounting for multiple testing (false discovery rate [FDR] ≤5%, horizontal dashed line). The vertical dashed line corresponds to a hazard ratio (HR) = 1. We labeled proteins of interest, including three members of the IL18 cascade. (**B**) Mortality HR in GEN-MOD (x-axis) and the UKBB (y-axis) are correlated for 1,421 proteins present in both datasets. For GEN-MOD and UKBB, the HR are per normalized standard deviation units of protein levels. We labeled in red proteins that have significantly different HR (heterogeneity FDR ≤5%) between GEN-MOD and UKBB. We provided protein name labels for seven proteins with a nominal heterogeneity P-value (HetP) <1×10^-4^. (**C**) Forest plot for 15 mortality-associated proteins with the smallest heterogeneity P-values between GEN-MOD and the UKBB. GALNT2 is a risk protein in GEN-MOD, but protective in the UKBB. DAPP1 is associated with increased mortality in GEN-MOD, but not significant in the UKBB.

In the UKBB, Gadd et al. analyzed 1,468 plasma proteins measured with Olink and found 652 proteins significantly associated with mortality^28^. While the UKBB and GEN-MOD are fundamentally different – the UKBB is a population-based cohort of mostly European-ancestry individuals not ascertained for a specific disease^29^ – we tested whether the same proteins associate with mortality in both cohorts. Of the 499 proteins significantly associated with mortality in GEN-MOD, 253 proteins were tested in the UKBB and 208 (84%) were associated with mortality with a consistent direction of effect (binomial P-value <2.2×10^-16^)(**Supplementary Table 9**). We found that the protein-mortality hazard ratios were correlated between GEN-MOD and the UKBB, but tended to be larger in GEN-MOD (**Fig. 2B-C**). However, this was not true for all proteins. For instance, GDF15, a protein strongly associated with mortality in the UKBB (HR=2.2, P-value <1×10^-300^) (ref. ^28^) and previously implicated in human longevity^30^ and chronic diseases^31–33^, was only marginally associated with mortality in SCD patients (HR=1.5, nominal P-value=0.020, FDR=0.10) (**Supplementary Table 9**). Together, our results suggest that a similar plasma proteome is associated with mortality in non-SCD participants and SCD patients, but the magnitude of the association of the proteins with survival is different, presumably because of the unique pathophysiological environment encountered in SCD (e.g. chronic hemolysis, systemic inflammation, high reactive oxygen species [ROS] production)^34^.

### Clinical variables and mortality-associated proteins

To better understand how the plasma proteome can predict SCD mortality, we tested in GEN-MOD the associations between the levels of the 499 mortality-associated proteins with 13 clinical parameters and five SCD-related complications (**Supplementary Tables 1** and **6**, all data collected at baseline). We found that increased levels of many of these proteins were positively correlated with WBC counts, reticulocyte counts, lactate dehydrogenase levels, dense RBC percentage and mean corpuscular hemoglobin (MCH), and negatively correlated with RBC counts, hemoglobin levels, hematocrit, HbF levels and eGFR (**Fig. 3A** and **Supplementary Tables 10-11**). We noted a unique pattern for platelet counts, with significant enrichments for both positively and negatively correlated proteins (**Fig. 3A** and **Supplementary Table 11**).

**Figure 3.**
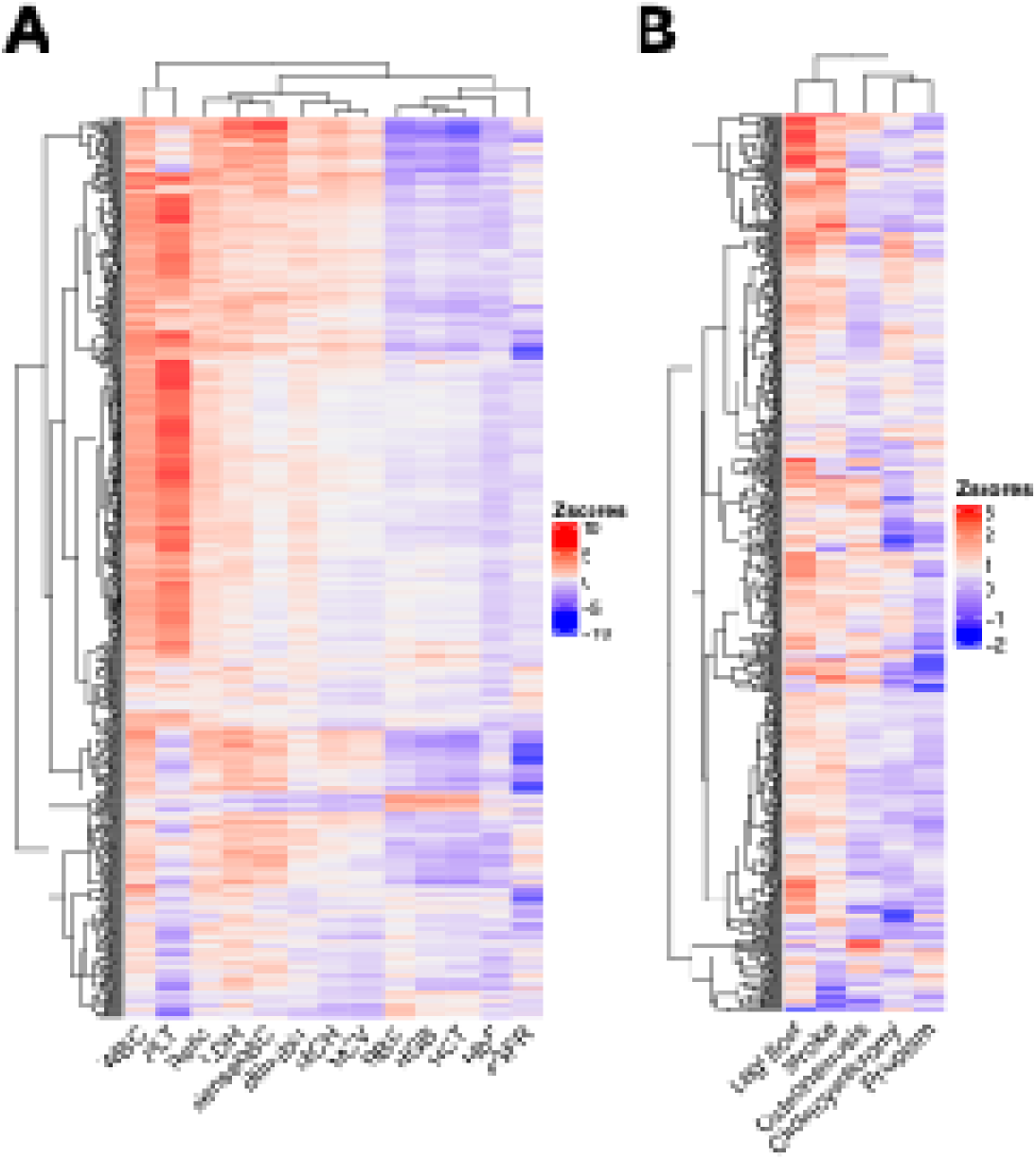
Associations between baseline clinical variables and mortality-associated plasma proteins. Associations between (**A**) 13 blood-based biomarkers routinely measured in SCD patients or (**B**) five SCD-related complications with the levels of 499 plasma proteins (y-axis) associated with mortality in GEN-MOD. We used |Z-score| ≥1.96 to define nominal significance. The red and blue colors indicate positive and negative associations, respectively. WBC, white blood cell; PLT, platelet; Retic, reticulocyte; LDH, lactate dehydrogenase; denseRBC, dense red blood cell; MCH, mean corpuscular hemoglobin; MCV, mean corpuscular volume; RBC, red blood cell; HGB, hemoglobin; HCT, hematocrit; HbF, fetal hemoglobin; eGFR, estimated glomerular filtration rate.

For the SCD complications, we observed an enrichment of mortality-associated proteins that are positively correlated with leg ulcers (**Fig. 3B** and **Supplementary Tables 10-11**). The causes of leg ulceration in SCD are not fully understood but likely involved vasculopathy defects and systemic inflammation^35^. Thus, the enrichment observed with leg ulcer may reflect the abundance of positive correlations between high mortality-associated protein levels and WBC counts (**Fig. 3**). It is interesting to note that while the presence of leg ulcers at baseline is not significantly associated with mortality in GEN-MOD (**Supplementary Table 6**), 107 proteins positively associated with leg ulcers are also strongly and positively associated with mortality prospectively (**Supplementary Tables 10-11**). This result highlights an advantage of an unbiased proteomic approach, which can nominate biomarkers that can predict future events (i.e. deaths) even in the absence of clear clinical manifestations at baseline.

### Developing a simpler protein signature of SCD mortality

The association between protein signatures developed in non-SCD populations and SCD mortality is promising, but we wandered if we could use the GEN-MOD plasma proteome data to develop a more SCD-specific signature of mortality. Ideally, this new signature would also include fewer proteins to facilitate future clinical implementation.

To prioritize a set of plasma proteins associated with survival in SCD patients, we applied a Cox Proportional-Hazards model with LASSO penalization (**Methods**). This approach identified five proteins (IL18R1, LGMN, MEX3B, MMP7, VEGFA), which were combined into a Cox LASSO mortality risk score associated with mortality (HR=5.70, P-value=1.8×10^-11^). With a concordant index (c-index) of 0.81, the model provided better mortality prediction than IL18R1 protein expression alone (c-index=0.76), the most significant protein associated with survival in GEN-MOD (**Supplementary Table 7**). In comparison, the c-index for the Conventional and Conventional adjusted models were 0.66 and 0.69, respectively (**Fig. 1C**). We validated the GEN-MOD-based Cox LASSO mortality score in the non-SCD UKBB cohort (4,401 events among 39,306 participants) using the expression of four of the five proteins available in this dataset (IL18R1, LGMN, MMP7, VEGFA). Applying the coefficients derived from the GEN-MOD cohort, this score demonstrated good predictive performance for mortality in the UKBB, with a c-index of 0.72. The mortality hazard ratio associated with this four-protein score was higher in GEN-MOD (HR=5.4, P-value= 2.8×10^-10^) than in the UKBB (HR=4.1, P-value<2.2×10^-16^). Importantly, we acknowledge that the performance of this simple four-protein signature is not immune to over-fitting issues and will require validation in external SCD cohorts with proteomic data available.

The four-protein mortality score was strongly associated with several clinical variables and complications in directions consistent with SCD pathophysiology: a significant negative association with eGFR (P-value=9.4×10^-12^), and positive associations with leg ulcers (P-value=1.6×10^-5^), dense RBC (P-value=1.3×10⁻) and osteonecrosis (P-value=2.4×10^-4^)(**Supplementary Table 12**). Notably, none of the four proteins used in the model were individually associated with leg ulcers or osteonecrosis at baseline. In addition, although the four proteins were not included in the mortality models from Goeminne et al. ^11^, the resulting four-protein signature was positively associated with mortality predictions from those models (**Supplementary Table 13**).

### The plasma proteome captures accelerated aging in SCD

Using different modalities (e.g. imaging, DNA methylation), previous studies have found that SCD patients age prematurely^36,37^. We calculated Conventional and organ-specific predicted biological age in GEN-MOD using plasma proteome-based models and compared it to chronological age (i.e. DeltaAge)(**Supplementary Table 4**)^11^. For the Conventional age model, the difference between biological and chronological age was larger in SCD patients (+6.0±5.4) than in non-SCD participants (+2.0±5.0) (**Fig. 4A**). We validated this result using an independent protein-based aging clock model (**Supplementary Fig. 5A**)^12^. For the organ-specific biological age models, we also observed that DeltaAge was larger in SCD patients when compared to non-SCD participants (**Fig. 4A**). Mortality in GEN-MOD was associated with DeltaAge calculated using the Skin and Intestine models, but not with the Conventional model (**Supplementary Fig. 5B**). DeltaAge across all models were not as strongly associated with baseline variables as the predicted mortality scores described above, with the exception of DeltaAge calculated with the Brain model that was strongly associated with stroke events that occurred retrospectively (**Fig. 4B** and **Supplementary Table 14**).

**Figure 4.**
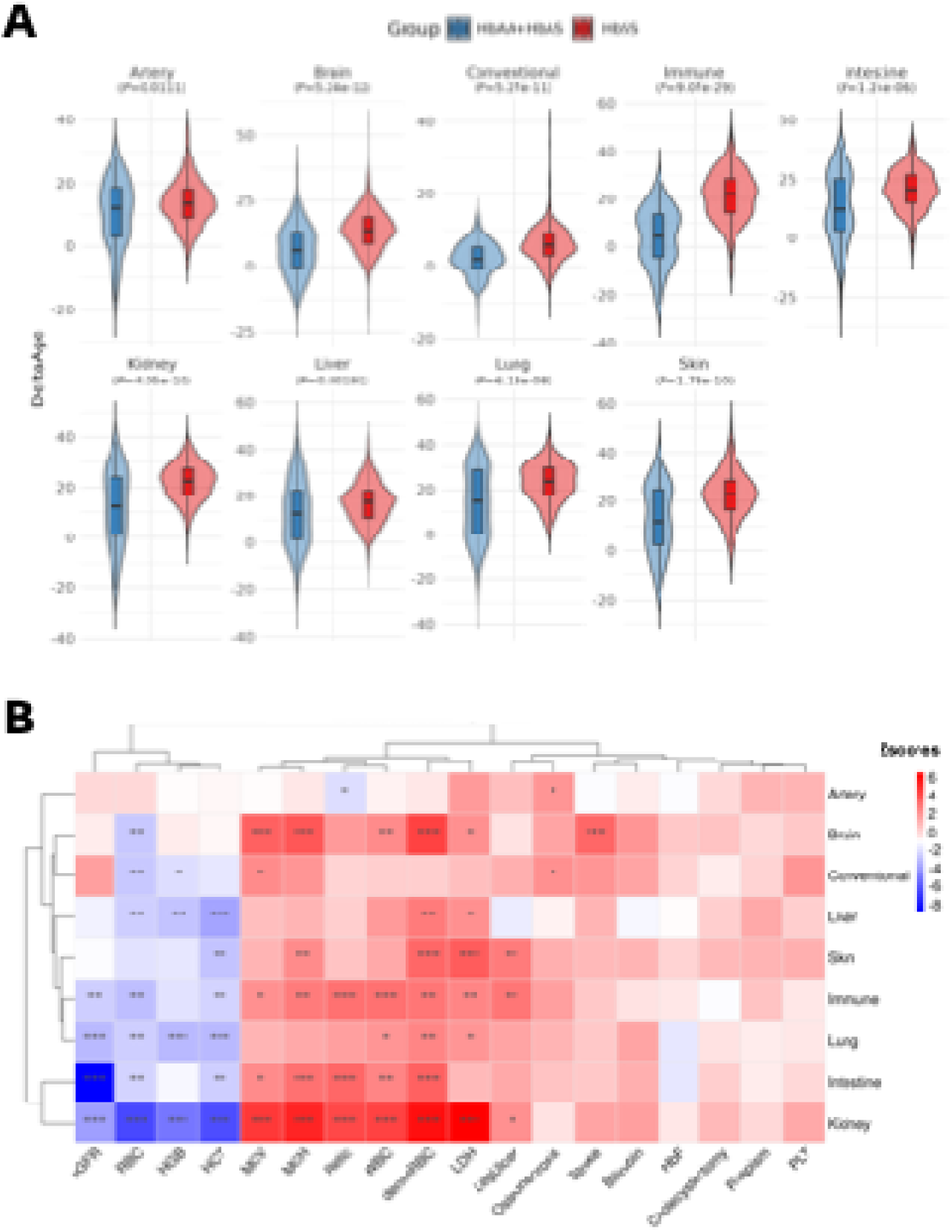
Plasma proteome-based biological clocks capture accelerated aging in SCD. (**A**) The difference between the predicted biological age calculated with the plasma proteome and chronological age (y-axis, DeltaAge) is larger in GEN-MOD SCD patients than in non-SCD participants from the same cohort. We calculated P-values using Wilcoxon’s test. (**B**) Heatmap that summarizes the association between DeltaAge from the Conventional or organ-specific models and baseline clinical variables. (*, nominal P-value <0.05; **false discovery rate <0.05; ***Bonferroni-corrected P-value <0.05; correction applied separately for each model). WBC, white blood cell; PLT, platelet; Retic, reticulocyte; LDH, lactate dehydrogenase; denseRBC, dense red blood cell; MCH, mean corpuscular hemoglobin; MCV, mean corpuscular volume; RBC, red blood cell; HGB, hemoglobin; HCT, hematocrit; HbF, fetal hemoglobin; eGFR, estimated glomerular filtration rate.

## DISCUSSION

One of the hallmarks of SCD is its clinical heterogeneity: patients with the same β-globin genotype can have mild, almost asymptomatic disease whereas others can suffer from life-threatening complications. While research has identified causal mechanisms (e.g. hemolysis, vaso-occlusion)^34^ and pinpointed some of the severity modifiers (e.g. HbF, α-thalassemia, pollution, social determinants such as access to healthcare services, patient stigmatization or even racism)^38–42^, it remains challenging to predict clinical outcomes in SCD. Identifying specific and sensitive biomarkers of SCD complications would greatly improve patient care. In this study, we showed that plasma proteome signatures developed in the non-SCD UKBB were predictive of mortality in the prospective GEN-MOD cohort. While associated with many relevant baseline clinical variables (e.g. WBC counts, dense RBC) and complications (e.g. leg ulcers), plasma proteome signatures outperform these variables in predicting SCD mortality.

It remains unclear how plasma proteome signatures can predict chronic diseases, biological age or mortality. A recent study suggested that these signatures mostly capture environmental risk factors and do not causally influence disease risk or mortality (preprint ^43^). Since the same signatures (**Fig. 1**) and a large extend of the same proteins (**Fig. 2-3**) predict mortality in the UKBB and GEN-MOD, it appears likely that the plasma proteome in SCD patients captures extrinsic factors that contribute to disease severity. Indeed, while the mortality proteome signatures were developed in the UKBB, they are correlated with dense RBC and leg ulcers (**Fig. 1E**), two characteristic features of SCD. Therefore, the plasma proteome signatures are biomarkers of general biological processes (e.g. increased systemic inflammation) that are influenced by the environment and predictive of disease onset or mortality. However, our results suggest that SCD creates a physiological environment that magnifies the predictive value of some of these proteins (**Fig. 2B-C**)

Our study has several limitations. First, we only measured proteins in the plasma of adult SCD patients such that our conclusions cannot be extended to pediatric SCD-related complications and mortality. Second, we did not analyze data from patients with other β-globin genotypes (e.g. HbSC, HbSβ^0^). Third, we did not assess the impact of current SCD treatments (e.g. hydroxyurea) on the plasma proteome; this will need to be evaluated in futures studies. Fourth, the predictive models that we used are likely suboptimal for SCD patients because they were developed in relatively healthy individuals of a mostly different ancestry (European-vs African-ancestry for SCD patients). Ideally, we would have access to proteomic datasets from large SCD cohorts to develop and calibrate SCD-specific predictive models, but such datasets are currently unavailable. However, using models developed in a completely different population offers the advantage to reduce the chance of data over-fitting and false positive results.

The implementation of using complex plasma proteome signatures in SCD clinics present with many technological and analytical challenges. For this reason, we explored the possibility of using a small number of proteins to predict SCD mortality. We found that a four-protein signature (IL18R1, LGMN, MMP7, VEGFA) could predict mortality in SCD patients and UKBB participants. It is critical to validate this simpler signature in external SCD cohorts.

In the clinic, we anticipate that plasma proteome signatures could be used together with existing strategies to identify adult SCD patients who would benefit from more frequent follow-up visits or more aggressive therapeutic interventions (e.g. cell or gene therapy). In the context of clinical trials, plasma proteome signatures could also be used as unbiased biomarkers to monitor the efficacy of new SCD treatments^44^. Overall, our results support plasma proteome signatures as new biomarkers of SCD severity and add proteomics to the toolkit to bring SCD one step closer to precision medicine.

## METHODS

### Study participants

Sample collections and procedures were in accordance with the institutional and national ethical standards of the responsible committees and proper informed consent was obtained. The project was approved by the Montreal Heart Institute Ethics Committee (project #09-1137).

### Plasma protein quantification

The Olink experiment was performed at the McGill Genome Centre. Plasma samples (40 µL) from 504 participants of the GEN-MOD cohort, including 396 SCD patients (HbSS) and 108 non-SCD individuals (56 HbAS and 52 HbAA), were analyzed using the Olink Explore 5K platform, which quantifies the expression of 5,416 proteins. To minimize batch effects, samples were randomized with the Well Plate Maker R package prior to transfer into 96-well Eppendorf twin.tec PCR plates.

### Quality control

Quality control was performed as described previously^10^. Briefly, proteins with ‘WARN’ or ‘FAIL’ QC flags were excluded before principal component analysis, resulting in the removal of APOE. Principal component one (PC1) and two (PC2) were used to identify outliers among participants and assays: points with values exceeding 5 standard deviations (SD) from the mean of PC1 or PC2 flagged two participants and three proteins (STK24, GAS8 and PHLDA3) as outliers. Additional filtering was performed using the OlinkAnalyze R package with interquartile range plots, identifying ten participant outliers. Missingness was evaluated for both participants and assays, and data with more than 11% missing values were excluded from downstream analyses, resulting in the removal of 14 participants and one protein (FN1). In total, 25 of 504 participants and 5 of 5,416 plasma proteins were excluded from subsequent analyses.

### Prediction of biological age and mortality

The predicted biological age and mortality risk for GEN-MOD participants were calculated using models developed by Goeminne et al^11^. In that study, plasma expression levels of 3,072 proteins measured with the Olink Explore platform in approximately 50,000 UK Biobank (UKBB) participants were used to develop organ-specific prediction models for 18 organs as well as a whole-organism model referred to as “Conventional”. We applied the coefficients provided in Supplementary Table S1A (for biological age) and S1C (for mortality) to GEN-MOD by summing the protein-specific coefficients from each organ model multiplied by their corresponding Olink-normalized expression values. For biological age predictions, intercept values of each model were added.

Because Olink provides relative quantification of protein expression, which can vary across platforms, we accounted for potential differences between the Olink Explore 5K platform used in GEN-MOD and the Olink Explore 3K platform used by Goeminne et al. by recalibrating the predictions using UKBB data. Specifically, we fitted separate linear regression models of predicted biological age or mortality risk on chronological age for GEN-MOD and UKBB. We then extracted the slopes and intercepts from each regression and recalibrated the GEN-MOD predictions as follows:

Recalibrated GEN-MOD predicted biological age or mortality risk = a + b × GEN-MOD predicted biological age or mortality risk,

where:

- b = slope for UKBB / slope for GEN-MOD
- a = intercept for UKBB − b × intercept for GEN-MOD

Finally, we evaluated the performance of each organ-specific biological age and mortality model in GEN-MOD by calculating the Pearson correlation coefficient (*r*) between predicted values and chronological age in non-SCD participants. Only organ models with *r*>0.3 and included by Goeminne et al.^11^ were retained for downstream analyses, resulting in the inclusion of eight organ models and the Conventional model. The DeltaAge for each organ was calculated as the difference between the predicted biological age and the chronological age. Comparisons between 376 SCD patients and 103 non-SCD participants were performed using the Wilcoxon test.

### Survival analyses

Survival analyses of SCD patients were conducted using Cox Proportional-Hazards regression models implemented in the survival R package. The analysis included 40 events (deaths) and 270 censored participants (controls). Associations between plasma proteins and survival were assessed using NPX values for each protein, which were rank-based inverse normalised and standardised to have a mean of and a SD of 1 prior to analysis, with models adjusted for chronological age and sex. For the association between predicted mortality and survival, the predicted mortality was also standardised to have a mean of 0 and SD of 1 before analysis. These models were adjusted either for chronological age and sex for all the organ models, and also for chronological age, sex, WBC, HbF, eGFR, and acute chest syndrome (coded as 0/1) for the Conventional model. The Kaplan–Meier survival curve was generated using the ggsurvplot function of the survminer R package, comparing the lowest (Q1) and highest (Q4) quartiles of the Conventional predicted mortality score. P-values were calculated with the log-rank test. Forest plots were generated with the forestplotter R package. We used the statistical method developed by Clogg et al. to assess the heterogeneity between the GEN-MOD and UKBB hazard ratios^45^.

### Associations with traits and complications

Associations of predicted mortality, DeltaAge, or plasma protein levels with clinical traits and complications in SCD patients were evaluated using regression models adjusted for chronological age and sex. Linear regression was applied to continuous variables (WBC, PLT, reticulocytes, LDH, dense RBC, bilirubin, MCH, MCV, RBC, HGB, HCT, HbF, and eGFR), while logistic regression was used for dichotomous outcomes (osteonecrosis, cholecystectomy, leg ulcer, priapism, and stroke, coded as 0/1). Z-scores for each association were calculated as the regression coefficient estimate divided by its standard error for heatmap visualization. Associations of predicted mortality or DeltaAge with the traits and complications were plotted using the pheatmap R package. We used the ComplexHeatmap R package to plot the data (unsupervised clustering) in **Figure 3**.

### LASSO-based Cox regression

Prioritization of a set of proteins associated with survival was performed using a Cox Proportional-Hazards model with LASSO penalization, implemented in the glmnet R package. Age and sex were included as unpenalized covariates, while penalization was applied to the protein variables. NPX values for each protein were rank-based inverse normalised and standardised to have a mean of 0 and a SD of 1 before analysis. The optimal penalty parameter (λ) was selected via 10-fold cross-validation. Survival analyses, including computation of the concordance index (c-index) and visualization of survival curves, were performed using the survival and survminer R packages. Replication in the UKBB cohort was performed by applying the coefficients derived from GEN-MOD for four of the five proteins in the model that were available in UKBB (IL18R1, LGMN, MMP7, VEGFA).

### Comparison with survival data in the UKBB

Comparisons of proteins associated with survival in GEN-MOD and UKBB were performed using data from Supplementary Table 4 of Gadd et al.^28^, which reported Cox Proportional-Hazards models for death adjusted for age and sex. Comparisons were restricted for proteins measured on both the Olink Explore 5K platform (GEN-MOD) and the Olink Explore 3K platform (UKBB).

### Differential expression analyses

Differential expression analyses of plasma proteins between SCD and non-SCD participants were performed using the limma R package. Linear models were fitted for each protein and chronological age and sex were included as covariates. P-values were corrected for multiple testing using the Benjamini–Hochberg false discovery rate method.

### Prediction of biological age using the ProtAge model

ProtAgeGaps were calculated using the model developed by Argentieri et *al*^12^. Of the 204 proteins in the published model, 196 were measured in the GEN-MOD cohort and included in this analysis. Protein expression values were normalized as described by the authors, and the pre-trained model was used to calculate protein-predicted ages (ProtAges). ProtAges in GEN-MOD were recalibrated using the ProtAges calculated in the UKBB, based on the slopes and intercepts of regression models of ProtAge on chronological age, as we described above. ProtAgeGaps were then defined as the difference between the recalibrated ProtAges and chronological age.

## ACKNOWLEDGMENTS

We thank all GEN-MOD participants who contributed data to this study. We also thank Pouria Jandaghi and Daniel Auld from the McGill Genome Center. We thank Austin Argentieri for help with the ProtAge model.

## ETHICS APPROVAL STATEMET

We collected data according to the Helsinki declaration and the study was approved by the Montreal Heart Institute ethics committee, Project #2009-106 (09-1137).

## DATA AVAILABILITY

The GEN-MOD proteomic data has not been deposited in a public repository because the data is not public but is available from the corresponding author on request. Scripts to analyze the data and draw figures are available at: http://www.mhi-humangenetics.org/en/resources/#anc_software.

## AUTHOR CONTRIBUTIONS

Conceived and designed the analyses: A.C. and G.L.; Collected the data: A.C., Y.Z., F.G. and P.B.; Contributed data: Y.Z., F.G. and P.B.; Performed analyses: A.C. and G.L.; Secured funding and supervised the work: G.L.; Wrote the manuscript: A.C. and G.L., with contributions from all authors.

