## Supplementary Figures for "Plasma proteome signatures are predictive of mortality in sickle cell disease"

**Supplementary Figure 1. Principal component analysis (PCA) of the proteomic dataset.** We ran PCA on 504 GEN-MOD participants and 5,416 plasma proteins (pre-quality control steps). The analysis included 396 HbSS, 56 HbAS and 52 HbAA participants. After data normalization and quality-control (QC), there is no batch effects due to technical plates (**A**) or biological sex (**B**, 1=Male, 2=Female). We note a separation of the participants due to the β-globin genotype on the PC1 axis: participants with the HbSS genotype (red) tend to be on the left of PC1 (**C**). The variance captured by PC1 and PC2 is 16.57% and 4.91%, respectively.

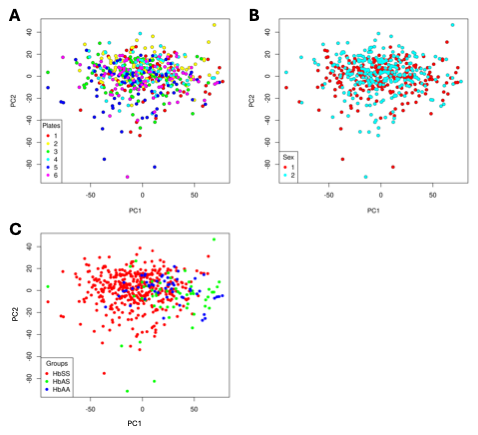

**Supplementary Figure 2. Many proteins are differentially expressed in the plasma of SCD vs. non-SCD participants.** Volcano plot that shows the differentially expressed proteins measured in the plasma of 376 SCD (HbSS) and 103 non-SCD (HbAS+HbAA) participants from GEN-MOD. The x- and y-axis represent the log_2_(fold-change) and -log_10_(FDR) values, respectively. The horizontal dashed line corresponds to the Benjamini-Hochberg corrected threshold of FDR=5% and the vertical dashed lines are at |log_2_(fold-change)|=1. L2FC, log_2_(fold-change); FDR, false discovery rate.

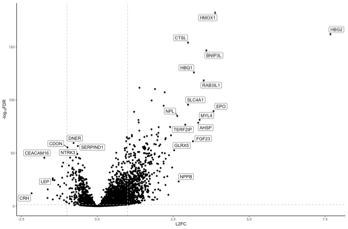

**Supplementary Figure 3.** Association between mortality and baseline clinical variables in GEN-MOD. We calculated hazard ratios (HR) using Cox Proportional-Hazards regression. None of the predictors are significantly associated with survival after accounting for multiple testing. See also **Supplementary Table 6**.

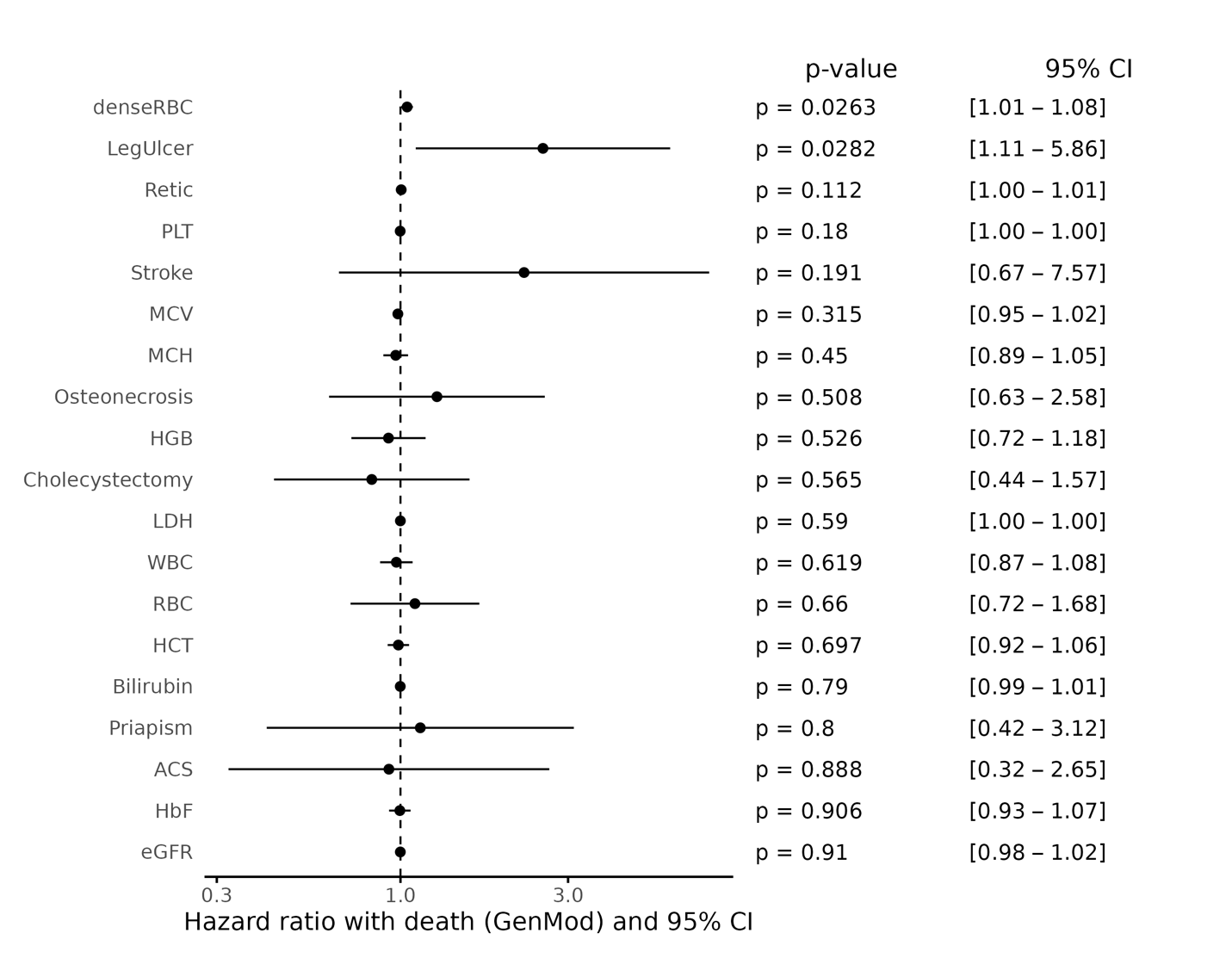

**Supplementary Figure 4. Organ-specific mortality prediction models.** (**A**) The predicted mortality scores calculated with organ-specific models^1^ are higher in SCD patients (HbSS) than in non-SCD (HbAS+HbAA) participants, except for the Artery model. (**B**) Using Cox Proportional-Hazards models, we tested the association between mortality in GEN-MOD SCD patients and organ-specific protein signatures of mortality. We corrected the models for biological sex and age at baseline. The forest plot shows the hazard ratio per standard deviation increase in the normalized prediction scores. After Bonferroni correction for eight tested models (α=0.00625), three models are significant: Lung, Skin and Artery.

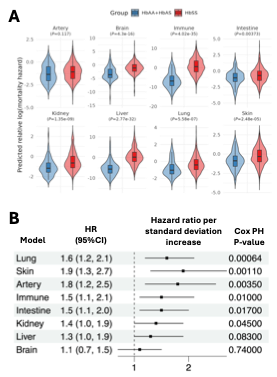

**Supplementary Figure 5. Biological age predictions using plasma proteome signatures.** (**A**) Using a different protein-based biological clock model^2^, we validated that the difference between biological and chronological age (y-axis, ProtAge) is larger in SCD than in non-SCD individuals. We calculated the P-value using the Wilcoxon test. (**B**) Using Cox Proportional-Hazards models, we tested the association between mortality in GEN-MOD SCD patients and DeltaAge calculated using previously described protein-based biological clock models^1^. We corrected the models for biological sex and age at baseline. The forest plot shows the hazard ratio per standard deviation increase in the normalized DeltaAge. After Bonferroni correction for nine tested models (α=0.0055), two models are significant: Intestine and Skin.

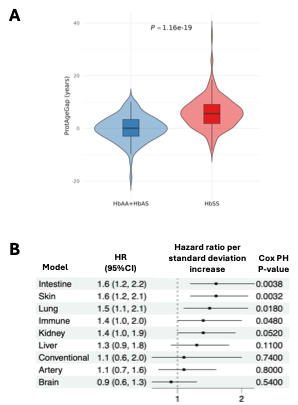
